# Impact of a city-wide school-located influenza vaccination program over four years on vaccination coverage, school absences, and laboratory-confirmed influenza: a prospective matched cohort study

**DOI:** 10.1101/19013151

**Authors:** Jade Benjamin-Chung, Benjamin F. Arnold, Chris J. Kennedy, Kunal Mishra, Nolan Pokpongkiat, Anna Nguyen, Wendy Jilek, Kate Holbrook, Erica Pan, Pam D. Kirley, Tanya Libby, Alan E. Hubbard, Arthur Reingold, John M. Colford

## Abstract

**Background:** It is estimated that vaccinating 50-70% of school-aged children for influenza can produce population-wide indirect effects. We evaluated a city-wide, school-located influenza vaccination (SLIV) intervention that aimed to increase influenza vaccination coverage. The intervention was implemented in over 95 pre-schools and elementary schools in northern California from 2014 to 2018. Using a matched prospective cohort design, we estimated intervention impacts on student influenza vaccination coverage, school absenteeism, and community-wide indirect effects on laboratory-confirmed influenza hospitalizations.

**Methods and Findings:** We used a multivariate matching algorithm to identify a nearby comparison school district with similar pre-intervention characteristics and matched schools in each district. To measure student influenza vaccination, we conducted cross-sectional surveys of student caregivers in 22 school pairs (2016 survey N = 6,070; 2017 survey N = 6,507). We estimated the incidence of laboratory-confirmed influenza hospitalization from 2011-2018 using surveillance data from school district zip codes. We analyzed student absenteeism data from 2011-2018 from each district (N = 42,487,816 student-days). To account for pre-intervention differences between districts, we estimated difference-in-differences (DID) in influenza hospitalization incidence and absenteeism rates using generalized linear and log-linear models with a population offset for incidence outcomes.

The number of students vaccinated by the SLIV intervention ranged from 7,502 to 10,106 (22-28% of eligible students) each year. During the intervention, influenza vaccination coverage among elementary students was 53-66% in the comparison district. Coverage was similar between the intervention and comparison districts in 2014-15 and 2015-16 and was significantly higher in the intervention site in 2016-17 (7% 95% CI 4, 11) and 2017-18 (11% 95% CI 7, 15). During seasons when vaccination coverage was higher among intervention schools and the vaccine was moderately effective, there was evidence of statistically significant indirect effects: adjusting for pre-intervention differences between districts, the reduction in influenza hospitalizations in the intervention site was 76 (95% CI 20, 133) in 2016-17 and 165 (95% CI 86, 243) in 2017-18 among non-elementary school aged individuals and 327 (5, 659) in 2016-17 and 715 (236, 1195) in 2017-18 among adults 65 years or older. The reduction in illness-related school absences during influenza season was 3,538 (95% CI 709, 6,366) in 2016-17 and 8,249 (95% CI 3,213, 13,285) in 2017-18. Limitations of this study include the use of an observational design, which may be subject to unmeasured confounding, and caregiver-reported vaccination status, which is subject to poor recall and low response rates.

**Conclusion:** A city-wide SLIV intervention in a large, diverse urban population decreased the incidence of laboratory-confirmed influenza hospitalization in all age groups and decreased illness-specific school absence rates among students during seasons when the vaccine was moderately effective, suggesting that the intervention produced indirect effects. Our findings suggest that in populations with moderately high background levels of influenza vaccination coverage, SLIV programs can further increase coverage and reduce influenza across communities.

## Introduction

Seasonal influenza contributes substantially to hospitalization and mortality, especially among infants and the elderly [1]. The economic burden of influenza as a result of lost earnings and medical cost was estimated to exceed $87 billion in the United States in 2007 [2]. To prevent the spread of influenza, seasonal influenza vaccination of all individuals over 6 months of age has been recommended by the Advisory Committee on Immunization Practices (ACIP) in the U.S. since 2010 [3]. Effectiveness of seasonal influenza vaccines varies from year to year depending on the quality of the influenza virus strain match and whether antigenic drift occurs between the time when the vaccine is manufactured and the start of the seasonal influenza epidemic, among other factors.

School aged children are responsible for the greatest proportion of community-wide influenza transmission, and as a result, efforts to increase vaccination are likely to have the largest impact on influenza transmission when targeted to them [4–9]. Mathematical models estimate that vaccinating at least 50-70% of school-aged children against influenza can prevent an influenza epidemic by producing herd immunity (i.e., “indirect effects”) [10,11]. In recent years, influenza vaccination coverage in the United States has ranged from 54-62% among elementary school-aged children and 37-44% among adults [12,13]. Healthy People 2020 set a goal of 70% influenza vaccination coverage for children and adults [14]. Mathematical models project that an increase in child influenza vaccination coverage to 80% would reduce influenza hospitalizations among children by 42% and among adults by approximately 20% [15]. Herd immunity is thought to play a critical role in reducing the burden of influenza morbidity and mortality among older adults, for whom vaccine effectiveness is frequently lower due to immunosenescence [16].

School-located influenza vaccination (SLIV) programs have been proposed as strategy to increase influenza vaccination coverage among children [16]. Prior studies reported that SLIV programs increased influenza vaccination [17–26] and decreased school absence [17–20,27–29] and student illnesses [17,20], and some studies report that their economic benefits likely outweigh the cost of program delivery [30,31]. There is some evidence that SLIV programs can produce community-wide indirect effects among pre-school aged children and adults; however, other studies have produced conflicting results [21,22,32–34]. Many prior SLIV evaluations did not use randomization or multivariate matching to select a comparison group and are subject to confounding [18,19,21,22,27–29,32–34]; those that have used more rigorous designs did not measure health outcomes [24–26,35,36] or enrolled small numbers of schools [17,20]. No prior studies have rigorously measured the impacts of large-scale SLIV interventions on student and community-wide health outcomes over multiple years.

Here, we report the findings of a four-year evaluation of a large-scale SLIV program in over 95 public, private, and charter schools in a diverse, urban, predominantly low-income city in northern California. The intervention was specifically designed to cover all public, private, and charter elementary schools in a geographical area in order to evaluate the potential for SLIV not only to reduce influenza among elementary schoolchildren but also to interrupt community-wide influenza transmission through herd effects. We expected that the SLIV intervention would reduce school absenteeism and community-wide influenza hospitalizations if it succeeded in increasing influenza vaccination coverage and if the seasonal influenza vaccine was at least moderately effective. Using a prospective matched cohort design and three independent data sources, we measured the intervention’s impact on student influenza vaccination and school absences and the incidence of community-wide laboratory-confirmed influenza hospitalization.

## Methods

### School-located influenza vaccination intervention

The Shoo the Flu intervention (www.shootheflu.org) delivered free influenza vaccinations at schools throughout the city of Oakland, California, including all of the public and charter elementary schools in Oakland Unified School District (OUSD, the “intervention district”) and offered delivery to all other charter, private, and pre-schools in Oakland prior to the start of influenza season from 2014 through 2018. OUSD enrolls a diverse, urban population of approximately 53,000 students, including over 26,000 elementary school students (grades K through 5). Over 70% of students in this district are from low-income households, and half of students speak a language other than English in their home. The intervention aimed to increase influenza vaccination coverage among primarily elementary school-aged children in order to reduce influenza among elementary schoolchildren and to produce indirect effects protecting other age groups in the community. In its first two years, the intervention deployed a mass media campaign in the Oakland area, including advertisements in the subway, bus shelters, billboards, and newspapers, as well as through digital media. The intervention did not carry out promotion efforts outside of the Oakland area, although it is possible that residents of areas near Oakland were exposed to Shoo the Flu media. Caregivers of the students provided written consent for vaccination, and this consent process was separate from consent to participate in the evaluation of the intervention. Children were eligible for vaccination regardless of their insurance status. From 2014-2018, between 95 and 138 elementary and pre-schools participated in Shoo the Flu, and each year the intervention vaccinated between 7,502 and 10,106 students (22-28% of eligible students) (S1 Appendix).

### Influenza vaccine effectiveness during the intervention

In 2014-15 and 2015-16, the intervention offered the live attenuated influenza vaccine (LAIV) to students and the inactivated injectable influenza vaccine (IIV) to students with LAIV contraindications, consistent with ACIP recommendations [37,38]. The intervention also offered the IIV to school staff and teachers. In early 2016 the ACIP changed its recommendation for children aged 2 to 8 years from LAIV to IIV due to concerns about the low effectiveness of LAIV in the two prior seasons [39]. In 2016-17 and 2017-18 the intervention offered only the IIV. The seasonal influenza vaccine delivered by the intervention had low effectiveness in 2014-15 and 2015-16 and moderate effectiveness in 2016-17 and 2017-18 (S2 Appendix) [40,41].

### Study design

The SLIV intervention was offered to all pre-schools and elementary schools in the city of Oakland with the goal of delivering SLIV to the largest number of schools possible in order to interrupt influenza transmission (i.e., produce indirect effects) in the city. For this reason, it was infeasible to use a cluster-randomized design because all schools in the district were offered the intervention. The study used a prospective matched cohort design to evaluate the SLIV intervention program [42]. We drew on multiple independent data sources to assess a full range of outcomes that could have been affected by the SLIV intervention: 1) influenza vaccination coverage, 2) influenza hospitalizations in the community, and 3) all-cause and illness-specific school absence rates among elementary school students. We conducted a survey of a sample of student caregivers to measure influenza vaccination coverage and analyzed existing school absence and influenza hospitalization records. In some cases, outcome assessment implied slightly different designs and estimators, as we describe below.

The matched design focused on the vaccination coverage survey. We first selected from comparison school districts among San Francisco Bay Area districts that had at least four elementary schools and had pre-intervention school-level characteristics similar to the intervention district. We restricted possible comparison districts to those with boundaries separated by at least five miles from the intervention district to minimize contamination. Though the intervention was provided to some private and charter schools, the study population was restricted to public elementary schools in the intervention city school district because pre-intervention data were not readily available to identify suitable comparison private or non-district charter schools.

We used a genetic multivariate matching algorithm [43] to pair-match schools in the intervention district with schools in each candidate comparison district. The matching algorithm used the following pre-intervention school-level characteristics: mean enrollment, class size, parental education, academic performance index scores, California standardized test scores, and the school-level percentage of English language learners and students receiving free lunch at school. We excluded pre-schools from this evaluation because the availability of pre-schools and enrollment criteria varied from school district to school district, complicating comparisons between districts. We identified West Contra Costa Unified School District (WCCUSD) as the best nearby comparison district because, on average, it had the smallest generalized Mahalanobis distance between paired schools [43] and a sufficient number of elementary schools (N=34 schools, grades K through 6) in the district to ensure adequate statistical power. The absolute value of the standardized difference was under 50 for most variables, indicating that the matching produced good quality school pair matches [44] (see S3 Appendix for further details).

Analyses of influenza hospitalization and school absences leveraged the matched design but used a slightly different approach tailored to each outcome. Because the intention was to measure community-wide indirect effects of SLIV on influenza hospitalization, we included hospitalizations of all residents of zip codes within the intervention and comparison district school catchment areas, including zip codes that were partially within the district boundary. To measure impacts on school absences we pre-specified inclusion of all public elementary schools enrolling Kindergarten to grade 5 (“K-5”) (50 intervention schools, 34 comparison schools) rather than the matched subset used to design the vaccine coverage survey in order to maximize precision.

### Outcomes and data sources

#### Vaccine coverage survey

We conducted two cross-sectional surveys of student caregivers to measure caregiver-reported student influenza vaccination, including vaccine type and vaccine provider. A survey in March 2017 measured vaccination from 2014-17, and a survey in March 2018 measured vaccination from 2017-18. We conducted the surveys in 22 of the 34 matched school pairs (22 K-5 schools in the intervention district and 22 K-6 schools in the comparison district). In all classrooms in each school, teachers distributed anonymous paper surveys to students to share with their caregivers. The survey was conducted independently from the intervention and allowed caregivers to report student influenza vaccination at any location. To estimate the required sample size for the survey, we assumed a realistic response rate of 50 surveys per school (approximately 16% of students), type I error = 0.05, intraclass correlation coefficient = 0.01, and 70% vaccine coverage in the comparison district. Enrolling 22 school pairs (44 schools total) allowed us to detect a minimum difference in vaccination coverage between districts of 6.5% with 80% statistical power.

#### Laboratory-confirmed influenza hospitalization

We obtained counts of all laboratory-confirmed influenza hospitalizations, intensive care unit admissions, and deaths among hospitalized patients and the duration of influenza hospitalization from zip codes within with the intervention and comparison school districts from 2011-2018 from the CDC-sponsored California Emerging Infections Program [45].

#### School absence records

We obtained records of absentee data for each student on each school day from 2011-2018 from all public elementary schools in each school district. Absences were classified by student grade, race/ethnicity, and absence type (all-cause absences versus illness-specific absences).

### Statistical analysis

Unless otherwise specified, analyses were conducted in R version 3.6.1 [46]. Our pre-analysis plan, selected datasets, and replication scripts are available through the Open Science Framework (https://osf.io/c8xuq/). We defined total effects as the difference in outcomes between elementary school-aged individuals (both those who did and did not participate in the intervention) in the intervention versus comparison site and indirect effects as the difference in outcomes between non-elementary school-aged individuals (individuals < 4 years, ≥13 years) in the intervention versus comparison site (Fig 1; for additional details on types of effects estimated, see S4 Appendix).

**Fig 1.**
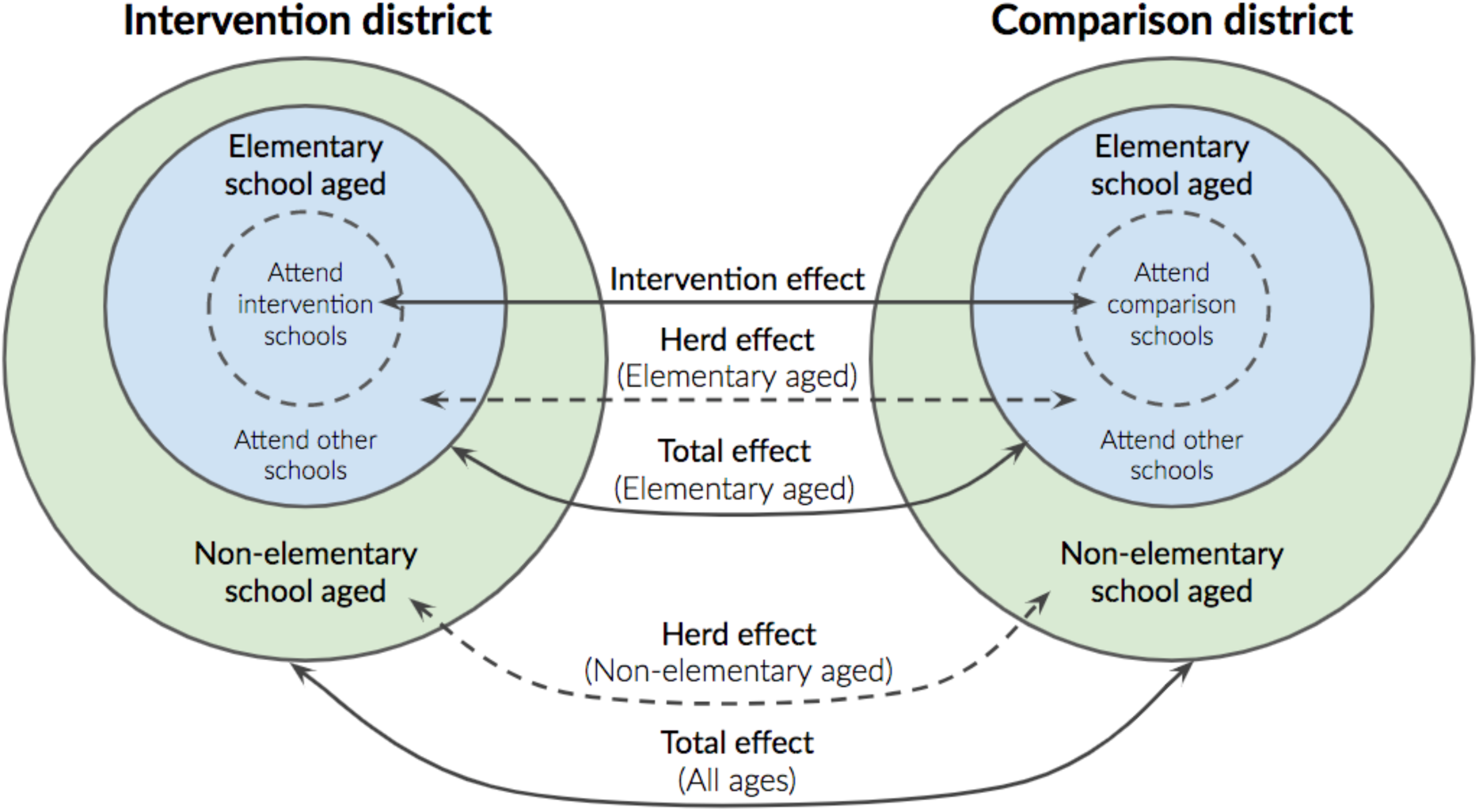
Schematic of total effects and herd effects estimated in this study. See additional intervention effect definitions in S4 Appendix.

#### Definitions of influenza season

Because influenza season timing varies from year to year, we pre-specified a data-derived definition of influenza season in order to conduct our analysis during the weeks in which the influenza epidemic occurred locally each year. Under this definition, influenza season started when there were at least two consecutive weeks in which the percentage of medical visits for influenza-like illness in California as reported by the California Department of Public Health [47] exceeded a cutoff, and the season ended when there were at least two consecutive weeks in which the percentage was less than or equal to a cutoff. We examined seasons defined using cutoffs of 2%, 2.5%, and 3% and selected 2.5% as the primary definition because it best captured seasonal variation in peak influenza-like illness (S5 Appendix). In addition, in a sensitivity analysis we used the California Emerging Infection Program’s surveillance definition of influenza season, which classifies influenza season as the weeks between October 1 and April 30 each year. This definition is widely used but is conservative in that it often includes many weeks of the year with limited influenza-like illness in influenza season.

#### Influenza vaccination coverage

We estimated influenza vaccination coverage and 95% confidence intervals using robust sandwich standard errors that accounted for clustering at the school level [48]. We estimated differences in vaccination coverage between districts using a generalized linear model that adjusted for student race/ethnicity and caregiver’s education level and estimated standard errors that accounted for clustering within matched school pairs. We restricted the analysis to grades K-5 because the intervention district’s elementary schools did not include 6^th^ grade. To assess possible selection bias among the sample of caregivers who responded to the survey, we also estimated vaccination coverage after standardizing the distributions of race/ethnicity and education in the sample to the pre-intervention percentages using data from the California Department of Education for the 44 participating schools and for the entire districts (see details in S6 Appendix).

#### Laboratory-confirmed influenza hospitalization

To estimate the cumulative incidence of influenza hospitalization, we obtained age-and race-specific population counts from the 2010 U.S. Census in the same set of zip codes used to identify influenza cases. We also obtained more recent annual population counts from the 5-year American Community Survey (ACS). Because counts were similar between both data sources and the ACS did not provide population counts for race and age in years, we used the U.S. Census data in our primary analyses. We fit log-linear Poisson models to estimate cumulative incidence using a log population offset and adjusting for age, sex, and race [49]. To control for pre-season differences between districts, we estimated the difference-in-differences (DID) defined as the difference in incidence in the intervention district prior to and during the intervention minus the difference in incidence in the comparison district prior to and during the intervention. Examination of pre-intervention absence trends indicated that the equal trends assumption was met (S7 Appendix). The DID parameter eliminates any time-invariant confounding and accounts for differences in pre-intervention trends between districts [50]. We also estimated the relative reduction adjusting for pre-intervention differences between districts, which we defined as 1 – RR x 100%, where RR is defined as the difference in incidence in the intervention district prior to and during the intervention divided by the difference in incidence in the comparison district prior to and during the intervention. We obtained standard errors for each quantity using the delta method. Consistent with our pre-analysis plan, we did not estimate DIDs for intensive care unit admission on its own or for influenza mortality because these are rare outcomes, and we were likely to be underpowered to detect an effect. We performed a sensitivity analysis using the following alternative influenza season cutoffs: 1) two consecutive weeks in which the percentage of medical visits for influenza-like illness exceeded 2% or 3% and 2) the period between week 40 and week 20 of each year.

#### School absence rates

We restricted the primary analysis of school absence rates to school days when both districts were in session. In addition, we restricted to school days that occurred during influenza season because we did not expect the intervention to influence influenza and absenteeism outside of that period. We also estimated outcomes in the peak week of influenza season, defined as the week with the highest proportion of influenza-like illness visits in California when school was in session. We estimated DIDs in mean absence rates using linear regression models and adjusted for available time-variant covariates: student race, grade, and month of absence. We did not adjust for pre-intervention school characteristics (i.e., those used in the matching of school pairs) because they were time-invariant and thus would have no effect on DID estimates. We calculated 95% confidence intervals using robust standard errors that accounted for clustering within schools [48]. We estimated the difference in total student absences during influenza season by multiplying DID estimates and confidence interval bounds by the total student enrollment and the number of school days in each influenza season.

To detect potential differential measurement error of school absences, we conducted a negative control analysis [51] using the school days in August, September, May, and June; these months were prior to the delivery of vaccines at school and outside of influenza season, when we did not expect to see an effect of the intervention [14]. We also performed a sensitivity analysis using the following alternative influenza season cutoffs: 1) two consecutive weeks in which the percentage of medical visits for influenza-like illness exceeded 2% or 3% and 2) the period between week 40 and week 20 of each year. We performed pre-specified subgroup analyses by student race, grade, and month.

As we describe in the Results, the negative control analysis indicated possible differential measurement error of school absences, so we performed a post-hoc probabilistic bias analysis to quantify the possible influence of outcome misclassification on our results [52]. We focused on outcome misclassification because exposure misclassification was highly unlikely, and our DID analysis accounted for measured and time-invariant unmeasured confounders. Through conversations with each school district, we defined distributions of the sensitivity and specificity of absence classification (S8 Appendix).

To examine impact across different levels of program participation among the 50 SLIV intervention schools, we predicted the mean absence rates (*Y*) setting each school’s value to each observed level of school participation in SLIV (*A*) and adjusting for school-level covariates student race/ethnicity, the percentage of students in each grade, average enrollment, mean class size, mean parent education level, percentage of English language learners, percentage of students receiving free lunch, mean 2012 Academic Performance Index score, mean 2013 Academic Performance Index score, and mean California Standards Test scores (*W*) (Σ_*w*_ E[*Y*|*A = a, W = w*] *P(W=w))*. We used an ensemble machine learning algorithm that flexibly adjusted for covariates correlated with the outcome (p-value < 0.1) in statistical models. The algorithm included the following estimation methods: the simple mean, main effects generalized linear models, stepwise logistic regression, Bayesian generalized linear models [53], generalized additive models [54], elastic net regression [55], random forest [56], and gradient boosting [57].

### Ethical statement

This study was approved by the Committee for the Protection of Human Subjects at the University of California, Berkeley (Protocols # 2014-01-5960 and 2016-12-9406). Caregivers of students in the influenza vaccine coverage survey received a letter from the school district describing the purpose of the survey and providing details about the optional and anonymous nature of the survey. A waiver of documented informed consent was necessary to carry out the survey because in two years of pilot surveys in which we requested documented informed consent, the complexity of consent forms contributed to very low response rates that prevented us from collecting a sufficiently large sample to estimate vaccination coverage.

## Results

### Influenza vaccine coverage

Measured pre-intervention characteristics were similar in populations residing in the catchment areas of the intervention and comparison schools (Table 1). In March 2017, field staff disseminated 8,121 surveys in 22 schools in OUSD and 10,054 surveys in 22 schools in WCCUSD (S9 Appendix). The response rates were 28% (N=2,246 surveys) in OUSD and 38% in WCCUSD (N=3,824). One school in OUSD withdrew, and we excluded its matched pair from analyses comparing the two districts. In March 2018, the same schools were invited to participate, and the response rates were similar. In each survey, 34-40% of respondents had a higher than high school level education, approximately 22-27% of respondents’ primarily language spoken at home was Spanish. The most common student race/ethnicity was Latino (36-41% in intervention; 50-51% in comparison), followed by Asian (22-25% in intervention; 16-17% in comparison) and Black / African American (16% in intervention; 10% in comparison).

**Table 1.** Pre-intervention characteristics of the population in school district catchment areas ^*a*^

| **Characteristic** | **Intervention**<br>**(95% CI)** | **Comparison**<br>**(95% CI)** |
| --- | --- | --- |
| Median household income (\$) | 51,849 (50,460, 53,238) | 61,596 (59,662, 63,530) |
| Households below the poverty level (%) | 21 (20, 22) | 15 (13, 16) |
| Highest education level (%) |  |  |
| Less than high school | 16 (15, 18) | 14 (12, 17) |
| High school graduate | 24 (21, 26) | 30 (25, 34) |
| Some college or Associate's | 46 (43, 48) | 50 (46, 55) |
| Bachelor's degree or higher | 15 (12, 17) | 6 (4, 8) |
| Children attending private vs. public schools (%) |  |  |
| Public kindergarten | 87 (81, 92) | 86 (80, 92) |
| Private kindergarten | 13 (8, 19) | 14 (8, 20) |
| Public grade 1-4 | 89 (86, 92) | 84 (79, 88) |
| Private grade 1-4 | 11 (8, 14) | 16 (12, 21) |
| Public grade 5-8 | 89 (86, 91) | 87 (83, 91) |
| Private grade 5-8 | 11 (9, 14) | 13 (9, 17) |
| Race (%) |  |  |
| White | 41 (40, 42) | 48 (47, 50) |
| Black or African American | 26 (25, 27) | 17 (16, 18) |
| American Indian and Alaska Native | 16 (16, 17) | 19 (18, 20) |
| Asian | 9 (8, 10) | 8 (7, 9) |
| Native Hawaiian and other Pacific Islander | 1 (0, 1) | 0 (0, 1) |
| Other races | 6 (6, 7) | 6 (5, 7) |
| Two or more races | 26 (25, 27) | 33 (32, 35) |
| Hispanic or Latino ethnicity (%) | 41 (40, 42) | 48 (47, 50) |

<sup>a</sup> Data from the 3-year 2013 American Community Survey subset by school district boundaries

Influenza vaccination coverage (from any location) among K-5 elementary students did not differ statistically between the intervention and comparison districts in the first two years of the SLIV intervention but was higher in the intervention district in the latter two years of the intervention. In relation to the comparison district, influenza vaccination coverage in the intervention district was 7% higher in 2016-17 and 11% higher in 2017-18; differences were statistically significant (Table 2). Standardizing vaccination coverage by student race and parent education produced similar results (S10 Appendix Fig 1). The percentage of elementary students vaccinated for influenza at school was 14% in 2014-15, 23% in 2015-16, 24% in 2016-17, and 26% in 2017-18 (S10 Appendix Fig 2). The majority of students not vaccinated at school were vaccinated at a doctor’s office or health clinic.

**Table 2.**
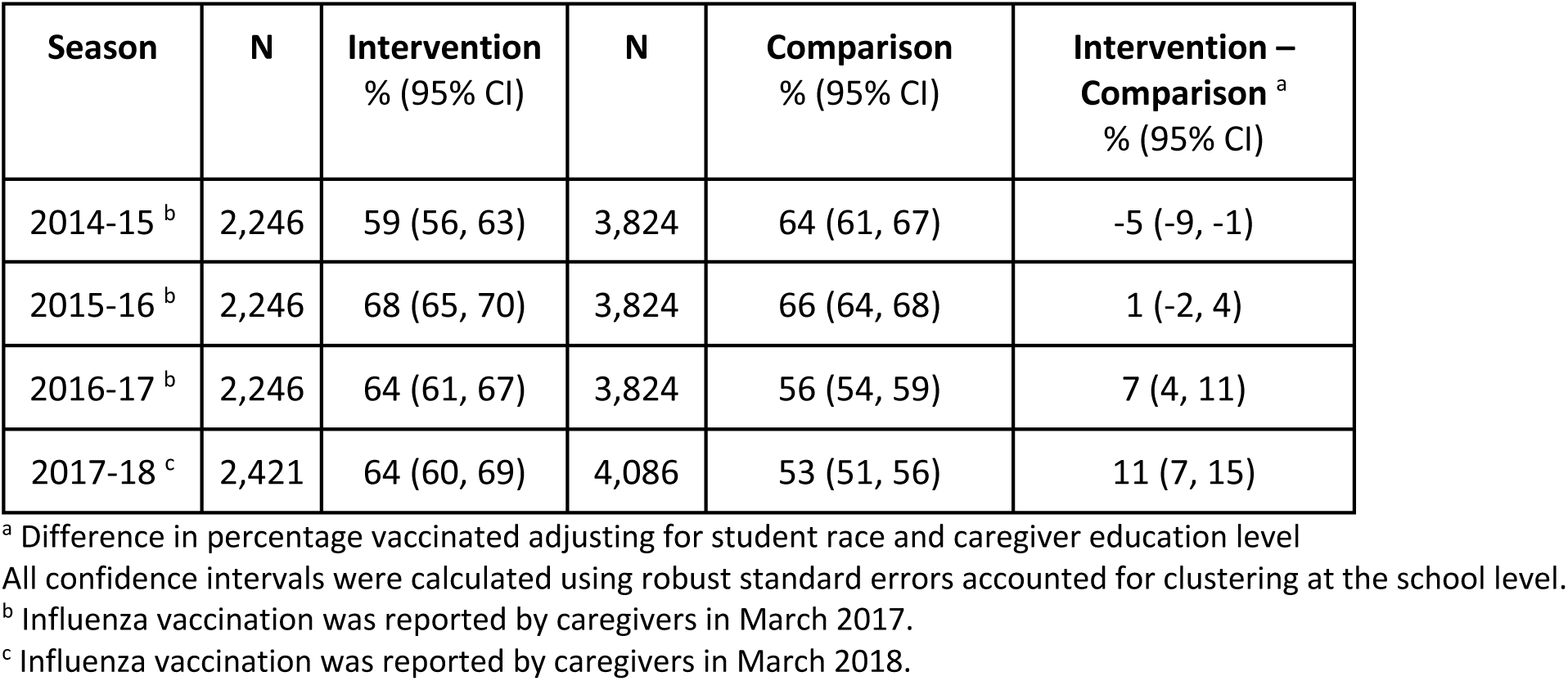
Caregiver-reported influenza vaccination coverage among elementary school students during the SILV intervention period

**Fig 2.**
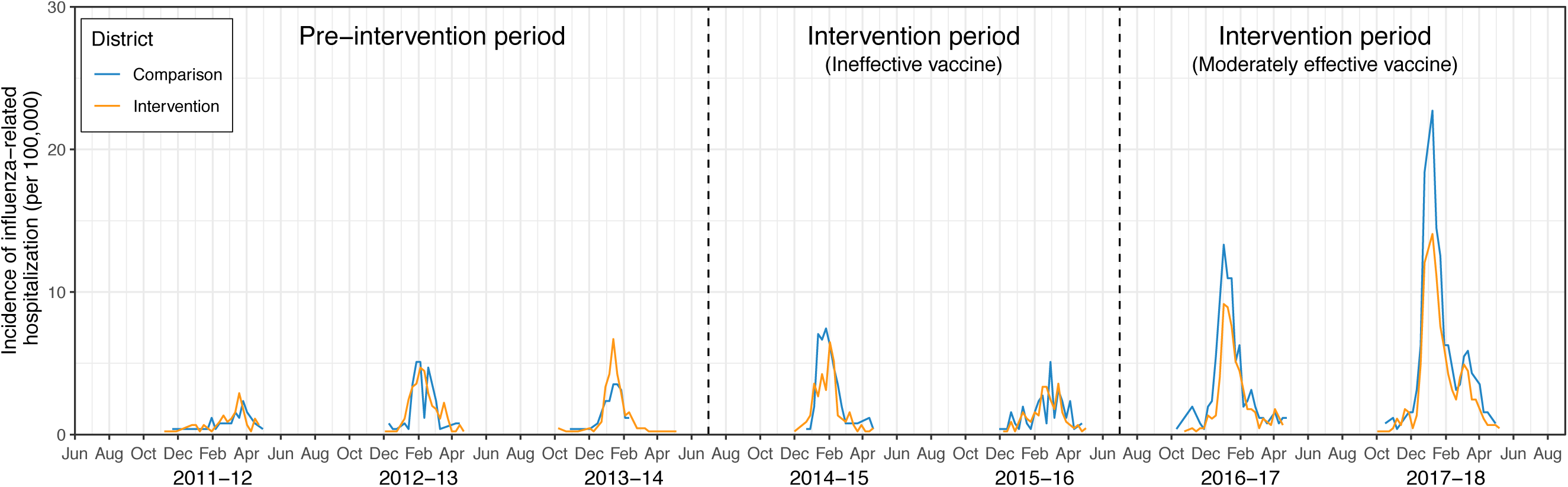
Weekly incidence of inpatient laboratory-confirmed influenza prior to and during the intervention. Weekly incidence proportion of laboratory-confirmed influenza hospitalizations between Week 40-52 and Week 1-20 of each year. Hospitalizations included school district residents tested at health care facility laboratories located in zip codes overlapping with OUSD and WCCUSD (Alameda County Public Health Department, Children’s Hospital Oakland, Contra Costa Public Health Department, Kaiser Permanente, Sutter Health). Population denominators were obtained from the U.S. 2010 Census using the same set of zip codes.

Influenza vaccination coverage varied by student race (S10 Appendix Fig 3). In 2016-17 and 2017-18, the highest coverage levels were among Asian and White students and the lowest were among Black / African American students and those whose race was not reported. Over the four years when the program was delivered, there were statistically significant increases in influenza vaccination coverage in the intervention district relative to the comparison district among Black / African American, Asian, Latino, and White students. The differences in coverage between districts was most striking among Latino students (12% higher in OUSD in 2016-17; 14% higher in 2017-18) and White students (10% higher in OUSD in 2016-17; 24% higher in 2017-18). The difference in student influenza vaccination coverage between districts was largest among students whose primary caregiver had a high school level education or less (S10 Appendix, Fig 4). When adjusting for school-level characteristics, influenza vaccination coverage did not vary by the percentage of students participating in SLIV in each school (S10 Appendix, Fig 5).

**Fig 3.**
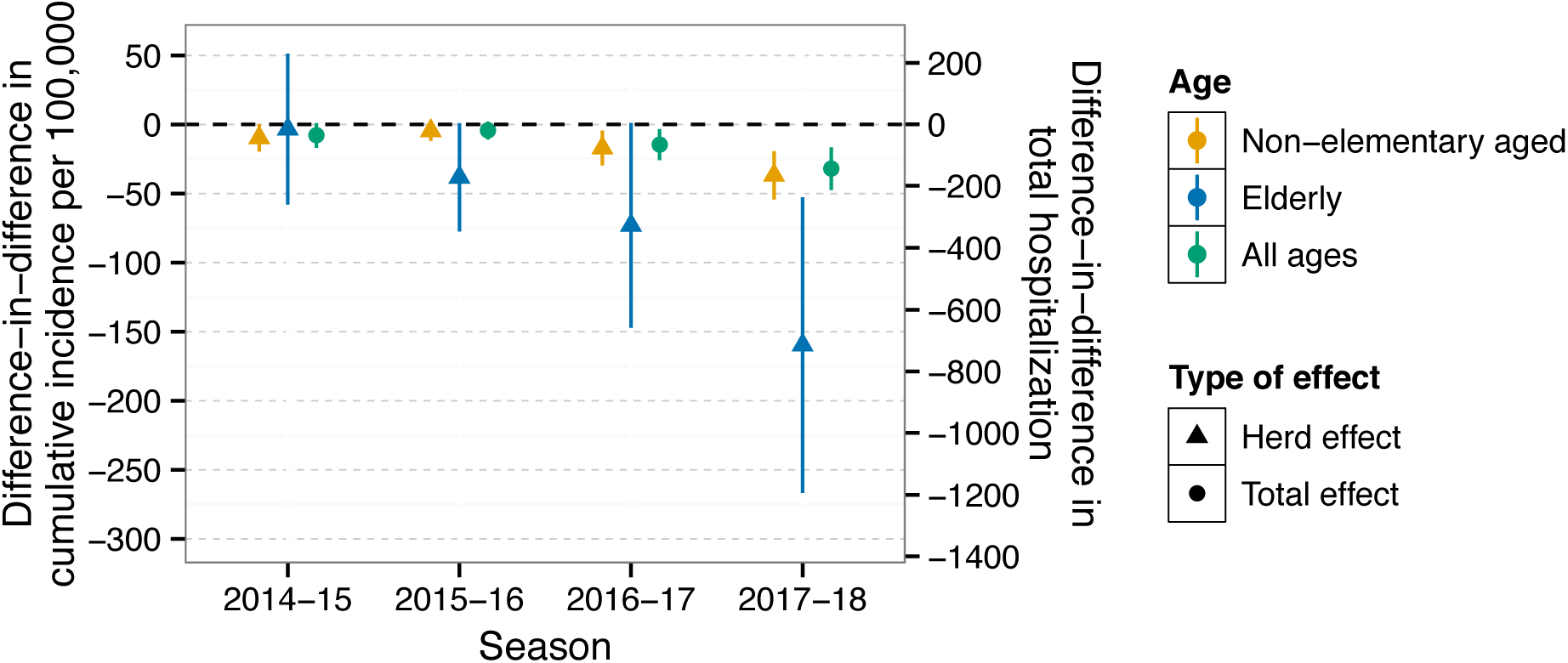
Total and indirect effects on cumulative incidence of inpatient laboratory-confirmed influenza during influenza season. Cumulative incidence of laboratory-confirmed influenza hospitalization and intensive care unit admission during influenza season. Difference-in-difference estimates represent the difference between intervention and control groups in their change in incidence from the three pre-program years (2011-2013) to each program year, which removes any time-invariant differences between groups (measured or unmeasured). The left y-axis presents the difference-in-difference in the cumulative incidence per 100,000. The right y-axis presents the difference-in-difference in the total hospitalizations, which was calculated as the product of the difference-in-difference in the cumulative incidence and the population of the intervention site. Parameters were estimated using a log-linear Poisson model with an offset for population size, and were further adjusted for age, race, and sex. Standard errors and 95% confidence intervals were obtained using the delta method.

**Fig 4.**
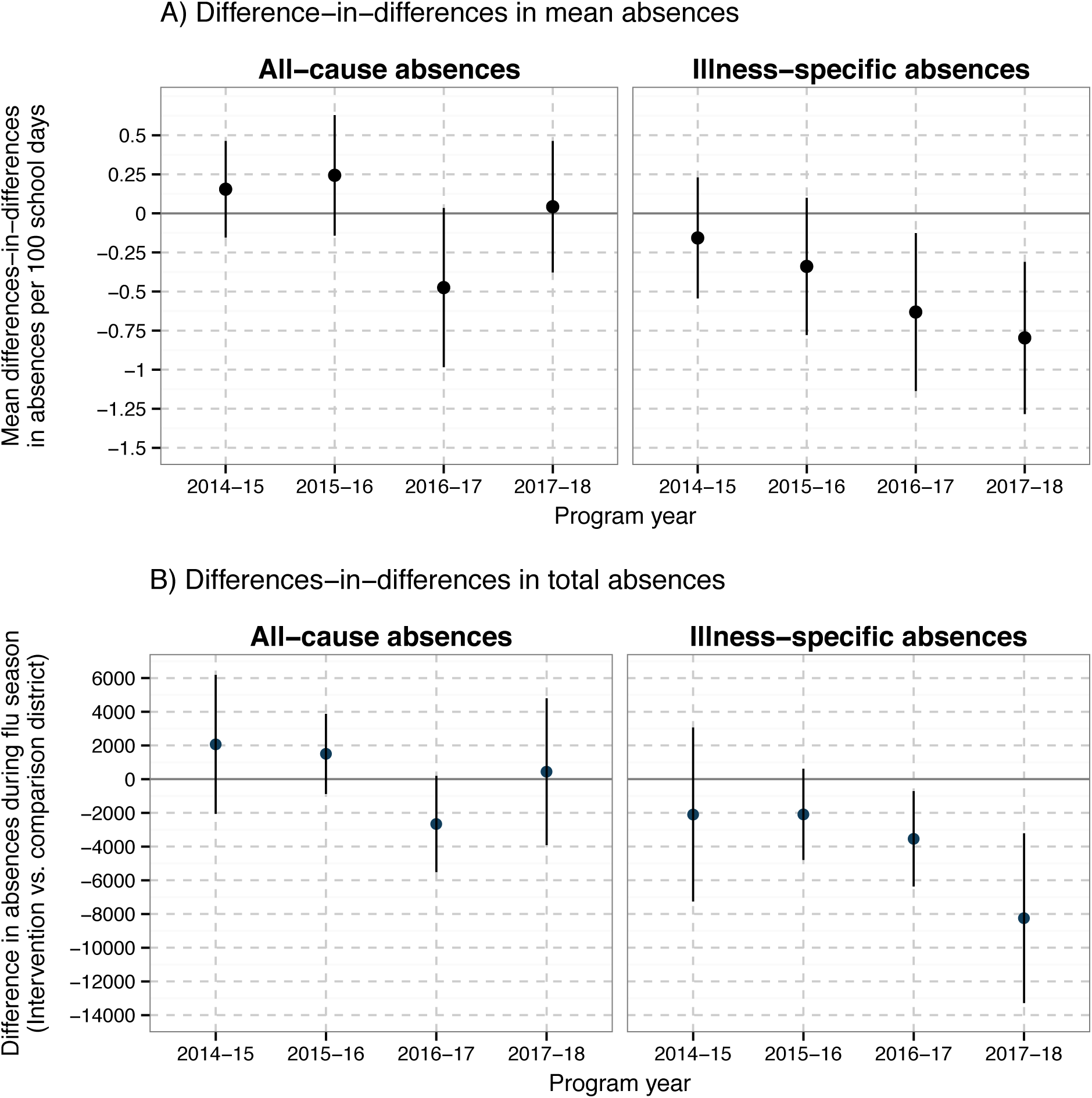
Intervention effects on the school absence rate per 100 school-days. In Panel A, each difference-in-difference estimate compares the difference in mean absence rates in each district in a program year compared to the three pre-program years (2011-2013); in Panel B, each difference-in-difference estimate compares the difference in total absence rates, which was calculated by multiplying difference-in-difference in mean absences by the total enrollment and total number of school days during influenza season each season. Difference-in-difference parameters remove any time-invariant differences between groups (measured or unmeasured). Parameters were estimated using a generalized linear model and were adjusted for month, student race, and grade. Standard errors and 95% confidence intervals account for clustering at the school level. Note: in 2011-12, 2016-17, and 2017-18, the peak week of the percentage of influenza-like illness visits in California were the last week of December, which coincided with school breaks, so for the absentee analysis we shifted the peak week definition to closest week with the next highest percentage of influenza-like illness visits when both school districts were in session.

### Influenza hospitalization

In the three years before the intervention, the incidence of influenza-related hospitalization was similar between the intervention and comparison districts (Fig 2). In 2014-15 and 2015-16, the incidence of influenza hospitalization was not statistically different between the intervention and comparison districts in any age group (Fig 3, S10 Appendix Table 1). In 2016-17 and 2017-18, hospitalization incidence was lower in the intervention versus comparison district in all age groups. Among non-elementary aged individuals (0-4, >14 years), the DID in total influenza hospitalization was −76 (95% CI −133, −20) in 2016-17 and −165 (95% CI −243, −86) in 2017-18 (Fig 3, S10 Appendix Table 1) in the intervention versus comparison district. Among individuals aged at least 65 years, the DID in total influenza hospitalization was −327 (95% CI −659, −5) in 2016-17 and −715 (95% CI −1,195, −236) in 2017-18 (Fig 3, S10 Appendix Table 1). Results were similar in analyses restricted to the peak week of influenza hospitalization (S10 Appendix Fig 6). When stratifying by patient race/ethnicity, in 2016-17 and 2017-18 DIDs were the largest among Asian / Pacific Islanders and Whites (S10 Appendix Fig 7).

Among all ages and across all four years, the mean DID in length of influenza hospitalization was approximately 1 to 2.5 days lower in the intervention district versus the comparison district (S10 Appendix Figs 8-9). The incidence of influenza-related intensive care unit admissions was lower in the intervention site than in the comparison site in 2014-17 and was similar in 2017-18 (S10 Appendix, Fig 10). The influenza-related mortality rate was slightly higher in the intervention site before the intervention and was lower in 2014-15, 2015-16, and 2017-18 influenza seasons (S10 Appendix, Fig 11). However, for all intensive care unit and mortality estimates, 95% confidence intervals for district-specific estimates overlapped. Our sensitivity analyses using alternative population denominators, influenza case definitions, and influenza season definitions yielded similar results overall (S11 Supplement, Figs 12-13).

### School absenteeism

In the three years prior to the Shoo the Flu program, during influenza season, the mean absence rate per 100 days in the intervention versus comparison district was 4.85 versus 5.84 for all-cause absences and 2.84 versus 2.81 for illness-specific absences (S10 Appendix Table 2). The DID in mean illness-specific absence rates per 100 days was −0.16 (95% CI −0.54, 0.23) in 2014-15 and −0.34 (95% CI −0.78, 0.10) in 2015-16 (Fig 4, S10 Appendix Table 2). In 2016-17 and 2017-18 the DID in illness-specific absence rates per 100 days was lower in the Intervention district compared to the comparison district (2016-17 DID −0.63 (95% CI −1.14, −0.13); 2017-18 DID −0.80 (95% CI −1.28, −0.31)). The reduction in total illness-specific student absences during influenza season was 3,538 (95% CI 709, 6,366) in 2016-17 and 8,249 (95% CI 3,213, 13,285) in 2017-18 in the intervention district. For all-cause absences, the DIDs during influenza season were not statistically significant in any of the years of the program. During the peak week of influenza, there was evidence of larger reductions in illness-specific absence rates in 2014-15, 2016-17, and 2017-18, and there was a significant reduction in all-cause absences in 2017-18 (S10 Appendix Table 2). Our sensitivity analyses using alternative influenza season definitions were consistent with the primary analysis (S10 Appendix Fig 14).

When stratifying by student race/ethnicity, there were statistically significant reductions in all-cause and illness-specific absences among Latino students in 2016-17 and 2017-18 and a smaller reduction in absences for Asian students in 2016-17 (S10 Appendix, Fig 15). For students of other race/ethnicities, DIDs were not statistically significant. Grade-specific patterns were similar overall (S10 Appendix, Fig 16). Mean absence rates were not associated with the percentage of students in each school that participated in Shoo the Flu in 2014-15 and 2015-16. In 2017-18 the school-level SLIV participation rate was associated with a modest reduction in the mean absence rate when adjusting for potential school-level confounders (S10 Appendix Fig 17).

We performed a negative control time period analysis, estimating DIDs outside influenza season, when we did not expect the intervention to affect absence rates. Overall, we did not see an effect on all-cause absences outside of influenza season. However, there were statistically significant reductions in illness-specific absences outside of influenza season in 2015-18 (S10 Appendix, Fig 18), suggesting that differential measurement error may have impacted the primary analysis. We explored the influence of outcome misclassification on our findings with probabilistic bias analysis under assumed distributions of sensitivity and specificity of outcome classification. We found that the majority of bias-corrected DID estimates in 2016-17 and 2017-18 indicated a reduction in both types of absences in the intervention district (S8 Appendix Figs 2-3). These findings suggest that outcome misclassification was not strong enough to alter the scientific inferences in our primary analysis of absenteeism.

## Discussion

Here, we evaluated the impact of a city-wide SLIV intervention delivered to over 95 elementary schools in a diverse, predominantly low-income city. During the first two years of SLIV, the program offered LAIV, which had low effectiveness [58,59], and on the whole, we did not observe impacts of SLIV in those years. In the latter two years, when the intervention delivered the IIV and the vaccine was moderately effective [41,60], we observed reductions in school absences and reductions in hospitalizations among age groups not targeted by SLIV, suggesting that the intervention produced herd effects. Unique strengths of our study design include the use of multivariate matching and a differences-in-differences approach to minimize systematic differences between the intervention and comparison sites and the pre-specification of our statistical analysis plan [61]. Evaluating SLIV over multiple years enabled us to examine the impact of SLIV in influenza seasons with different levels of vaccine effectiveness, vaccine recommendations, and circulating strains of influenza. Furthermore, this study leveraged three distinct, independent data sources that provided internally consistent results.

The SLIV intervention was associated with increases in influenza vaccination coverage of up to 11 percentage points among elementary school students in the intervention site versus the comparison site. This increase in vaccination coverage is smaller than those reported in prior SLIV studies, which ranged from 7 to 41 percentage points. However, in most prior studies, coverage at baseline or in the comparison group was substantially lower than 50% [17,18,20,22,24–26,34], while in our study it was 53-66%. It may be more difficult for SLIV to substantially increase coverage at moderate pre-intervention coverage levels. In addition, the switch from LAIV to IIV in the third year of the Shoo the Flu intervention may have inhibited larger increases in coverage because many children and/or caregivers prefer the nasal spray generally and in a school setting, and media coverage of poor LAIV effectiveness may have increased vaccine hesitancy for all influenza vaccine formulations. Vaccination coverage in the intervention district grew relative to the comparison district over time, in part because the comparison district coverage did not recover from the decline associated with the switch from LAIV to IIV vaccines. It is possible that coverage in the intervention district vaccine coverage will continue to grow in future years as the intervention builds trust and recognition.

A key question is whether SLIV interventions increase vaccination coverage among students who would otherwise not be vaccinated, whether they merely shift vaccination location from health care providers to schools, or whether both occur. Interventions that vaccinate children who would otherwise not have been vaccinated will have the largest impact on influenza transmission. Our findings suggest that both phenomena occurred in this study. Coverage was higher in the intervention versus comparison district in the final two years of the evaluation, suggesting that some otherwise unvaccinated children were vaccinated by the SLIV intervention. In addition, the proportion of students vaccinated at school in the intervention district (24-26% in the latter two years (S10 Appendix, Fig 2)) exceeded the difference in coverage between districts (7-11%), suggesting that approximately 15% of students vaccinated by the SLIV intervention would otherwise have been vaccinated through other means. A limitation is that pre-intervention vaccination data were not available, so we were not able to conduct DID analyses to account for any pre-SLIV differences in vaccination coverage between districts.

Influenza vaccination coverage in the Shoo the Flu intervention site was 64%, which was up to 11% higher than in the comparison site and well within the 50-70% range in which herd immunity is expected [10,11]. We observed 36-60% relative reductions in influenza hospitalization among non-elementary school aged community-members in seasons with moderate vaccine effectiveness (S10 Appendix Table 1). Indirect effects were strongest among individuals aged 65 years or older – the age group most vulnerable to influenza hospitalization and mortality. The magnitude of indirect effects we observed is similar to those of other SLIV interventions [21,22,32–34] and of interventions vaccinating children at any location [62]. In addition, our results are highly consistent with mathematical models, which project that an increase from 40 to 60% coverage in children aged 6 months to 18 years would reduce influenza hospitalization among adults over 19 years by 33-36% [15]. Our findings suggest that even modest increases in vaccination (i.e., up to 11%) associated with SLIV can produce meaningful community-wide reductions in influenza hospitalization, consistent with mathematical models [63].

We observed greater increases in influenza vaccination coverage in the intervention site versus the comparison site in students whose caregivers had up to a high school-level education compared to those with at least an associate degree. No prior SLIV evaluations have reported variation in the impact of SLIV on vaccination coverage by caregiver education level. Our findings contrast with those of prior studies of influenza vaccination outside school settings, which have generally found that higher education levels are associated with higher vaccination rates [64–66]. A limitation of this work is that education level was not available at the individual level for the influenza hospitalization and absenteeism outcomes.

Shoo the Flu’s impacts on influenza vaccination coverage, school absence rates, and influenza hospitalization varied by race/ethnicity. Black / African American students had the lowest influenza vaccination coverage, consistent with national CDC estimates [12], and a smaller increase in coverage in the SLIV district compared to most other races. This pattern in vaccination coverage may explain the more modest impacts on absenteeism and hospitalization among Black / African Americans compared to Whites and Asians. For Latino / Hispanic students, vaccination coverage was lower, but the increase in coverage in the intervention site was larger over time compared to White students. There was a larger reduction in absenteeism but a smaller reduction in hospitalizations in Latino / Hispanics compared to Whites and Asians. Disparities in influenza vaccination coverage may result from barriers to health care, differing knowledge and attitudes about vaccines, unconscious bias on behalf of providers promoting vaccination, underinsurance, or other social determinants of health [67]. Latino / Hispanics more commonly cite limited health care access and cost as barriers while Black / African Americans report mistrust in influenza vaccination [68]. In principle, SLIV should reduce disparities by race / ethnicity by increasing access to free influenza vaccination regardless of insurance status. However, increasing access alone does not address mistrust or vaccine hesitancy; to reduce disparities in vaccination, future interventions may need to couple SLIV with outreach strategies to address vaccine hesitancy, mistrust, and misconceptions about the influenza vaccine in particular [69]; incentives for vaccination; or policies mandating vaccination.

This analysis is subject to several limitations. First, although the reductions in illness-specific school absences we observed were of a similar magnitude to those reported in prior studies [17–20,27–29], our finding of significant differences in absence rates outside of influenza season suggests that our absentee results may be subject to differential misclassification. It is possible that school year- and school district-specific differences unrelated to the SLIV intervention could explain these findings. For example, district-specific policies to decrease absences around school breaks (some of which coincide with influenza season) or at the beginning or the end of the school year (which is included in our negative control time period analysis) may have impacted our estimates in an unknown direction, especially if they differed before and during the SLIV intervention. In addition, parents may have attributed illness absences to a different reason, and such misclassification could have varied by district. Our probabilistic bias correction analysis suggested that correcting for misclassification would not have changed our conclusion that the SLIV intervention reduced illness-specific absences in 2016-17 and 2017-18.

Second, given that the SLIV intervention was delivered city-wide, it was not possible to conduct a randomized trial. The matched cohort design minimized differences in measured confounders between the intervention and comparison site, and DID analyses controlled for measured and unmeasured time-invariant confounding. Nevertheless, unmeasured time-dependent confounding could still bias this observational design. For example, the first year of the SLIV intervention coincided with the rollout of the preventive benefits of the Affordable Care Act, which may have jointly affected health care utilization and vaccination patterns in the study region.

Third, our vaccination coverage estimates relied on caregiver reporting, which is subject to inaccurate recall and low response rates. Prior studies report that caregiver recall of child influenza vaccination in the past season has a sensitivity of 88-92% and a specificity of 82-90% compared to medical records [70–72]. Coverage estimates for 2014-15 and 2015-16 may be more vulnerable to measurement error because they rely on a 2-3 year recall period. Overall, our vaccination coverage estimates were consistent with caregiver-reported national and California-specific estimates from the Centers for Disease Control and Prevention [12,73].

Finally, we did not have a direct measure of laboratory-confirmed influenza incidence in elementary school children or influenza vaccination coverage estimates among non-elementary aged individuals; thus, it remains possible that factors other than the SLIV intervention could explain our findings. We were not able to link individuals between different data sources because personal identifiers were not available to us. Nevertheless, high levels of internal consistency across results from three independent data sources lends credence to the validity of our findings.

## Conclusions

Offering school-located influenza vaccination to all elementary schools in a large, urban district led to 7-11% increases in vaccination, which translated to meaningful reductions in community-wide reductions in influenza hospitalization and illness-specific absences among school children among those not targeted by the program, including the elderly. Our findings suggest that even modest increases in influenza vaccination above moderate coverage levels can produce broad community-wide benefits.

## Data Availability

Data from the vaccine coverage survey are available at https://osf.io/c8xuq/. Absentee data cannot be shared publicly because of protections under the Family Educational Rights and Privacy Act. Influenza hospitalization data may be requested from the California Emerging Infections Program.

## Acknowledgements

We would like to thank Aleta Rattanasith for her data collection and management, Hannah Warren for her consultation on community engagement and contribution to data collection, and Catharine Ratto and Amy Pine for technical advising as well as our partners at Shoo the Flu, the Alameda County Public Health Department, Oakland Unified School District, West Contra Costa Unified School District, California Emerging Infections Program, and Applied Survey Research for their contributions to this evaluation. This study was funded by the Flu Lab.

## Supporting Information Captions

S1 Appendix. Influenza vaccines delivered by the SLIV intervention each year

S2 Appendix. Estimated influenza vaccine effectiveness from 2014-2018

S3 Appendix. Selection of schools for the vaccine coverage survey

S4 Appendix. Definition of types of effects

S5 Appendix. Alternative influenza season definitions

S6 Appendix. Assessment of selection bias in the vaccine coverage survey

S7 Appendix. Pre-intervention trends in absences in each school district

S8 Appendix. Quantitative bias analysis to assess misclassification of absence rates

S9 Appendix. Vaccine coverage survey participant flow

S10 Appendix. Additional tables and figures

